# An Integrated Framework with Machine Learning and Radiomics for Accurate and Rapid Early Diagnosis of COVID-19 from Chest X-ray

**DOI:** 10.1101/2020.10.01.20205146

**Authors:** Mahbubunnabi Tamal, Maha Alshammari, Meernah Alabdullah, Rana Hourani, Hossain Abu Alola, Tarek M. Hegazi

## Abstract

Early diagnosis of COVID-19 is considered the first key action to prevent spread of the virus. Currently, reverse transcription-polymerase chain reaction (RT-PCR) is considered as a gold standard point-of-care diagnostic tool. However, several limitations of RT-PCR have been identified, e.g., low sensitivity, cost, long delay in getting results and the need of a professional technician to collect samples. On the other hand, chest X-ray (CXR) is routinely used as a cost-effective diagnostic test for diagnosis and monitoring different respiratory abnormalities and is currently being used as a discriminating tool for COVID-19. However, visual assessment of CXR is not able to distinguish COVID-19 from other lung conditions. Several machine learning algorithms have been proposed to detect COVID-19 directly from CXR images with reasonably good accuracy on a data set that was randomly split into two subsets for training and test. Since these methods require a huge number of images for training, data augmentation with geometric transformation was applied to increase the number of images. It is highly likely that the images of the same patients are present in both the training and test sets resulting in higher accuracies in detection of COVID-19. It is, therefore, vital to assess the performance of COVID-19 detection algorithm on an independent data set with different degrees of the disease before being employed for clinical settings. On the other hand, machine learning techniques that depend on handcrafted features extraction and selection approaches can be trained with smaller data set. The features can also be analyzed separately for various lung conditions. Radiomics features are such kind of handcrafted features that represent heterogeneous appearance of the lung on CXR quantitatively and can be used to distinguish COVID-19 from other lung conditions. Based on this hypothesis, a machine learning based technique is proposed here that is trained on a set of suitable radiomics features (71 features) to detect COVID-19. It is found that Support Vector Machine (SVM) and Ensemble Bagging Model Trees (EBM) trained on these 71 radiomics features can distinguish between COVID-19 and other diseases with an overall sensitivity of 99.6% and 87.8% and specificity of 85% and 97% respectively. Though the performance is comparable for both methods, EBM is more robust across severity levels. Severity, in this case, was scored between 0 to 4 by two experienced radiologists for each lung segment of each CXR image represents the degree of severity of the disease. For the case of 0 severity, sensitivity and specificity of the EBM method are 91.7% and 100% respectively indicating that there are certain radiomics pattern that are not visibly distinguishable. Since the proposed method does not require any manual intervention (e.g., sample collection etc.), it can be integrated with any standard X-ray reporting system to be used as an efficient, cost-effective and rapid early diagnosis device. It can also be deployed in places where quick results of the COVID-19 test are required, e.g., airports, seaports, hospitals, health clinics, etc.

## INTRODUCTION

COVID-19 is currently a major health crisis that the world is experiencing [1]-[2]. According to the World Health Organization (WHO) the disease is highly infectious and around 1 out of every 5 people who gets COVID-19 caused by 2019 novel coronavirus (2019-nCoV) needs to be hospitalized due to breathing difficulty [3]. To avoid burden on the healthcare system and to reduce the spread of the disease, it is vital to carry out fast and accurate diagnosis of COVID1-9. In clinical practice, real time polymerase chain reaction (RT-PCR) is considered as gold standard in COVID-19 diagnosis. Though the specificity of RT-PCR test is high, the sensitivity varies between 71 to 98% depending on sample collection site and sample quality [4-6], stage of disease [7] and degree of viral multiplication or clearance [8]. Other different types of samples (e.g., blood, urine, stool etc.) were also used for COVID-19 detection with variable results [9].

It has been reported that chest CT demonstrates higher sensitivity for the diagnosis of COVID-19 compared to initial RT-PCR tests of pharyngeal swab samples [10]-[11]. On the other hand, about 81% of the patients with negative RT-PCR results confirmed positive via chest CT scans [12]. According to the Radiological Society of North America (RSNA), the sensitivity of CT to detect COVID-19 infection was 98% compared to RT-PCR sensitivity of 71% [13]. Thus, CT could be considered as a primary tool for COVID-19 detection, unless the patient cannot be moved [14]. On the other hand, chest X-ray (CXR) has been considered as an insensitive method specially in the detection of COVID-19 at early disease. Because of lower spatial resolution compared to CT, CXR has a reported low sensitivity of 59% for initial detection and may appear normal until 4-5 days after the start of symptoms [15, 16]. Because of availability and portability, it can be utilized as both baseline and follow up imaging method for monitoring disease progression [17]. Other than availability and portability, CXR has several advantages over other conventional COVID-19 diagnostic tests, e.g., short examination time, low cost and can be sterilized easily and quickly [18].

Several deep neural network based machine learning approaches were proposed to detect COVID-19 directly from CXR. The reported sensitivity and specificity vary from 85.35 to 96.7% and 90 to 100% respectively based on methods, disease types and classification of the diseases [10, 19-22]. The detail of each method is provided in supplementary Table 1. To obviate the challenges of limited CXR COVID-19 data, transfer learning along with data augmentation was implemented which allows to increase the diversity of the data. However, it is to be noted that data augmentation can only increase the data via geometric transformation of the images and cannot increase the number of images with different COVID-19 conditions. The features that make appearance of COVID-19 different from other lung conditions are also not well understood with these methods.

**Table 1:** Training data description

| Disease Type | Description | No of CXR | Source |
| --- | --- | --- | --- |
| **non-COVID-19**<br>(Total 152 images) | Normal | 50 | JSRT [27] |
|  | Viral/bacterial pneumonia | 50 | Kaggle [28] |
|  | Other lung conditions (ARDS (4), SARS (15), Pneumocystis (12), Streptococcus (13), Chlamydomphila (1), E. Coli (4), Klebsiella (1) and Legionella (1)) | 52 | GitHub [29] |
| **COVID-19** | COVID-19 | 226 | GitHub [29] |

On the other hand, machine learning techniques that depend on handcrafted features extraction and selection approaches can be trained with smaller data set. The features can also be analyzed separately for various lung conditions. In depth analysis of different first, second and third order statistical features known as radiomics have been successfully utilized for decoding the radiographic phenotype in cancer [23, 24]. Recent studies prove the potential of differentiating glioblastomas (GBM) from metastatic brain tumors (MBTs) on contrast-enhanced T1 weighted imaging with radiomics based machine learning method [25]. However, only one study so far has reported to achieve 93% sensitivity and 90% specificity in detecting COVID-19 by applying different machine learning algorithm on textural features extracted from CXR [26].

The training and performance evaluation of all these methods were carried out by randomly splitting the data into training, validation and test sets. The performance of the method was not assessed on an independent CXR data set. On the other hand, the sensitivity and specificity of diagnosis of COVID-19 from CXR were not separately evaluated at different levels of severity of the diseases.

In this study, we are proposing a radiomics based machine learning approach to detect COVID-19 from CXR. The sensitivity and specificity of the method was verified on a completely independent test data containing both normal, viral and bacterial pneumonia and confirmed COVID-19 with different levels of severity determined by two experienced radiologists.

## MATERIALS AND METHODS

### TRIAING DATA SET AND RADIOMICS FEATURES EXTRACTION

In the proposed approach, each lung is first manually delineated from training CXR image set. The training dataset was created from three different available data repository containing normal and different types of lung conditions. Since the objective is to detect COVID-19 from other lung conditions, all CXR images were grouped into two classes (COVID-19 and non-COVID-19). The details of the training data is provided in Table 1

To avoid bias in class distribution for training, Adaptive Synthetic (ADASYN) oversampling approach was implemented on non COVID-19 dataset to balance the imbalanced dataset. ADASYN synthetically creates new samples in between difficult-to-classify samples from the minority class [30]. For the manual segmentation of lung on all the CXR images except the normal ones, MATLAB Image Segmenter app was used [31]. For the normal cases, the available lung masks were used [39]. 100 radiomics features were then extracted using the segmented lung and PyRadiomics tool for each lung separately [23]. This yielded 18 first-order statistics, 9 2D shape-based, 22 Gray Level Co-occurrence Matrix (GLCM), 16 Gray Level Run Length Matrix (GLRLM), 16 Gray Level Size Zone Matrix (GLSZM), 5 Neighboring Gray Tone Difference Matrix (NGTDM) and 14 Gray Level Dependence Matrix (GLDM) features.

### RADIOMICS FEATURES SELECTION

To select the features that have better classification ability between COVID-19 and others, two methods were implemented. The first method is a heatmap of Z-scores to identify features that can classify these two groups. The second method is the one-way ANOVA test to find the features that have a statistically significant difference between the means of the two classes with the criteria p<0.05. It was found that only 71 features out of 100 radiomics features show statistically significant difference and therefore, these features were only used for training the machine learning algorithms. Out of these 71 features 13 are first order,, 3 2D shape based, 20 GLCM, 8 GLDM, 10 GLRLM, 13 GLSZM and 4 NGTDM extracted features.

### TRAINGING OF MACHINE LEARNING ALGORITHMS

Number of supervised machine learning algorithms were evaluated using the Classification Learner App of MATLAB. To optimize the parameter of the machine learning algorithms to classify between COVID-19 and other cases, 10 fold cross validation approach was implemented. The overall process is shown in Figure 1.

**Figure 1:**
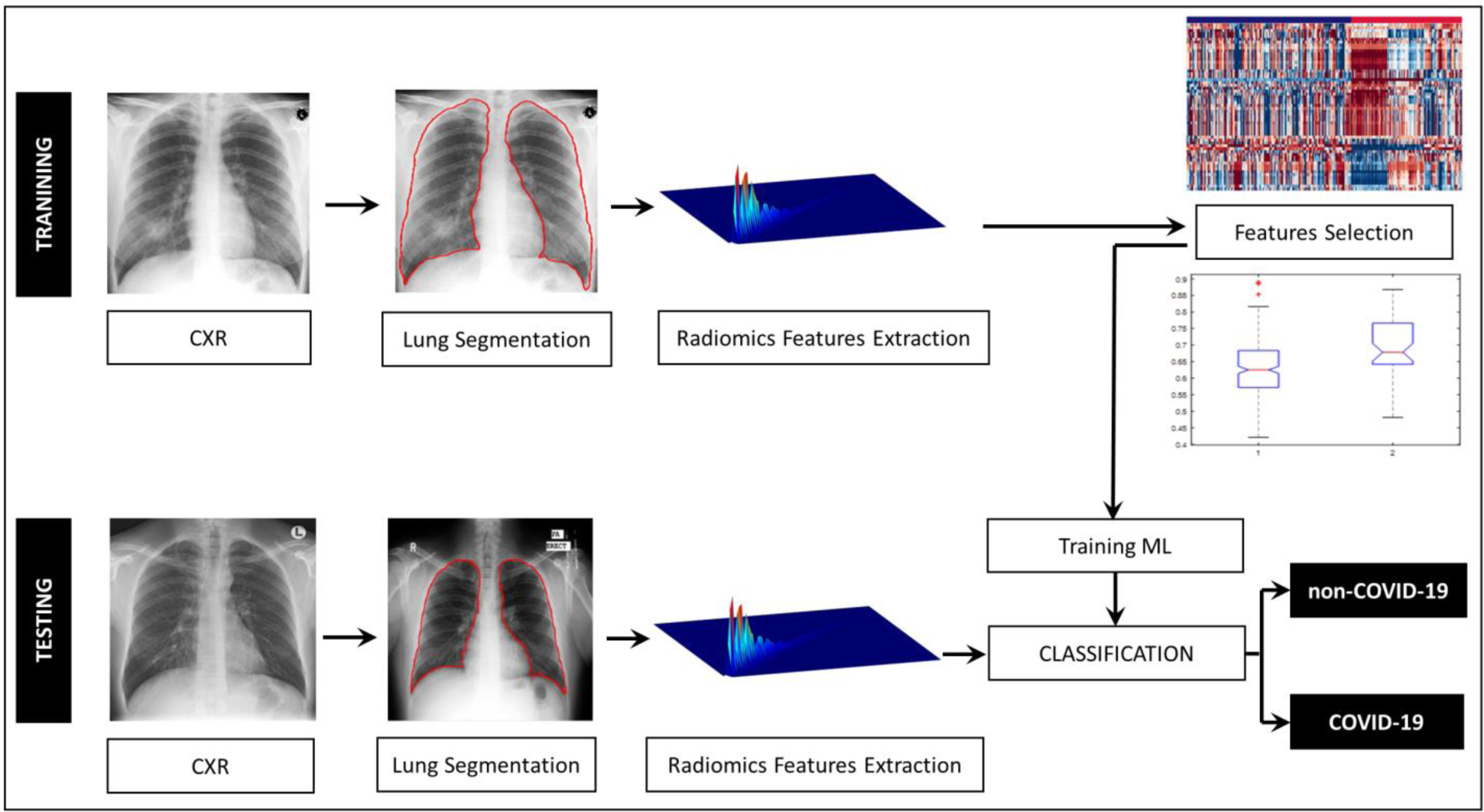
Illustration of the methodology for COVID-19 detection from CXR images.

Area under receiver operating characteristic (AUC-ROC) curve as well as sensitivity, specificity, and accuracy were calculated to evaluate the ability of the classifier to discriminate the COVID-19 CXR cases from the other cases. Sensitivity, specificity and accuracy were calculated as: 

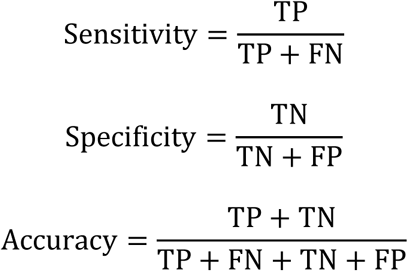

The three best performing classifiers during the training phase - 1) fine Gaussian support vector machine (SVM), 2) fine k-nearest neighbor (KNN) and 3) ensemble bagged model (EBM) trees were chosen for further evaluation on the test data.

### TEST DATA SET

Test CXR data set used in this study is an independent data set and consists of 165 CXR images (330 lungs) containing 25 normal, 25 viral/bacterial pneumonia and 115 COVID-19 cases. The 115 COVID-19 CXR images of 25 patients were acquired at the King Fahd Hospital of the University (KFHU), Imam Abdulrahman Bin Faisal University, Dammam, Saudi Arabia. COVID-19 was confirmed with standard RT-PCR test. The details of the test data is shown in Table 2.

**Table 2:**
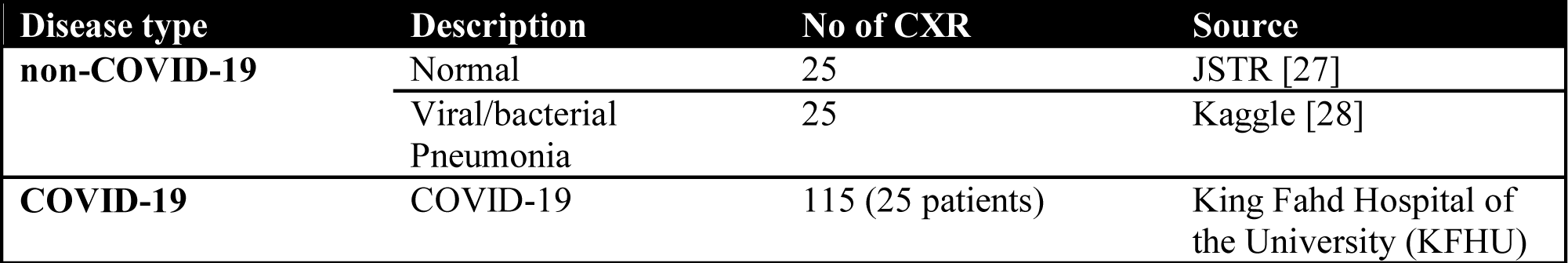
Test data description

Multiple chest X-ray were taken for all the 25 patients. Out of 25 patients 15 patients passed away and 10 patients were released cured from the hospital. Different levels of severity have been observed on each of the different CXR image of the same patient. Even for the same CXR, the severity between the two lungs are sometimes different according to the visual assessment of the radiologists. To evaluate the robustness of machine learning algorithms, the severity of each lung of each CXR for both COVID-19 and viral/bacterial pneumonia was scored in a scale of 0 to 4 based on the consensus of two experienced radiologists. The score 0 to 4 was assigned to each lung depending on the visual assessment on the extent of involvement by consolidation or GGO (0 = no involvement (clear lung), 1 = <25%, 2 = 25-50%, 3 = 50-75% and 4 = >75% involvement). Test data set based on severity is shown in Table 3. This is to be noted that during training severity was not taken in to consideration.

**Table 3:** Number of segmented lungs for each severity

| **Severity** | **0** | **1** | **2** | **3** | **4** | **Total** |
| --- | --- | --- | --- | --- | --- | --- |
| **non-COVID-19** | 50 | 17 | 22 | 11 | 0 | 100 |
| **COVID-19** | 36 | 88 | 71 | 27 | 8 | 230 |
| **Total** | 86 | 105 | 93 | 38 | 8 | 330 |

### VALIDATION ON TEST DATA SET

Similar to the training data set, 100 radiomics features were first extracted from each lung of each CXR image belonging to the test data set. Before extraction of the features, lungs were segmented manually on CRX. However, only 71 features that showed statistically significant difference between COVID-19 and other cases during training were used for performance evaluation. The three best performing classifiers during training were then applied using these 71 features to evaluate the classification performance of the machine learning algorithms. The performance was also evaluated on each separate lung to investigate the effects of severity on the classification performance. The same sensitivity, specificity and accuracy along with AUC-ROC were used for the performance evaluation on the independent test set.

## RESULTS

The list of the 71 radiomics features that are statistically significantly different between COVID-19 and non-COVID-19 CXR images are provided in supplementary Table 2 along with the p-value. Figure 2 shows the Z-score heatmap of the significant radiomics features for each CXR image ordered according to the diagnosis class. The selected features clearly display the difference between the COVID-19 and other cases.

**Figure 2:**
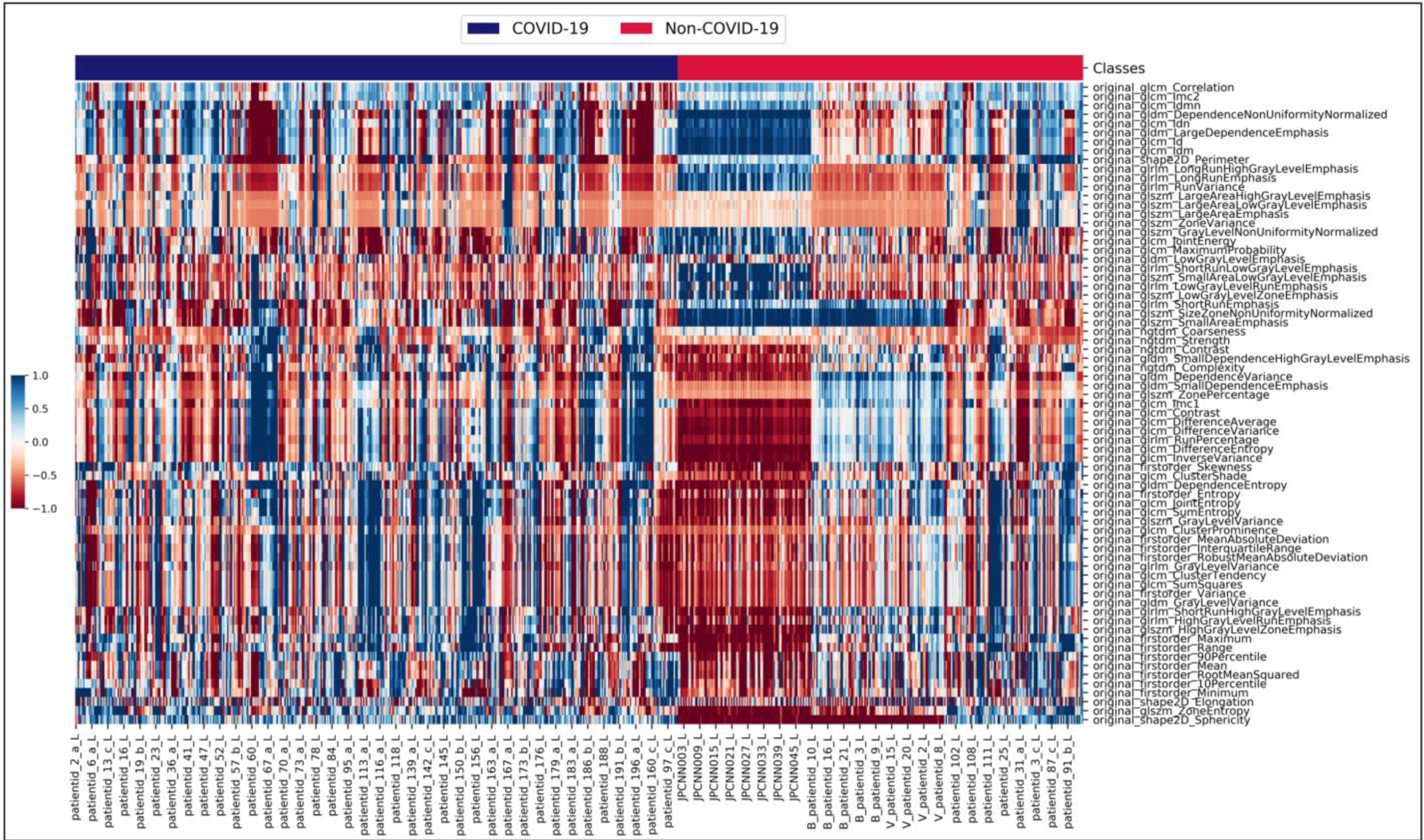
Z-score heatmap of 71 radiomics features that yield statistically significant difference between COVID-19 and others. Each row represents one feature and each column represents one CXR image used in the training set.

The performance of the classifiers during training is shown in Table 4. Each performance value shown in the table is the average of the 10-fold cross-validation results. From the table, we can see that the SVM classifier has the highest average sensitivity and AUC-ROC. But the lowest specificity. The highest specificity is achieved by fine KNN with 97.9% but with the lowest average sensitivity 88.9% and AUC-ROC of 0.9343. The sensitivity, specificity and AUC-ROC for EBM method is always more than 90%.

**Table4:** Performance of the classifiers during training

| Classifier | Sensitivity | Specificity | Accuracy | AUC-ROC |
| --- | --- | --- | --- | --- |
| Fine Gaussian SVM | 98.2% | 88.4% | 93.4% | 0.9894 |
| Fine KNN | 88.9% | 97.9% | 93.3% | 0.9343 |
| Ensemble Bagged Model Trees (EBM) | 91.6% | 92.6% | 91.8% | 0.9772 |

Table 5 shows the performance results of applying the previously trained classifiers to an independent test data set. The SVM classifier was able to correctly predict all the COVID-19 cases except one with 99.6% sensitivity, while EBM correctly predicted COVID-19 with 87.8% sensitivity. The sensitivity of the fine KNN was the lowest with 73.5%. On the other hand, the two highest specificity of 98% and 97% were achieved by KNN and EBM respectively, while the specificity of the SVM method was the lowest with 85%. The highest accuracy and AUC-ROC were achieved by SVM and EBM respectively (95.2% and 0.9241). Figure 3 shows the AUC of all the three machine learning algorithm during training and testing phases.

**Table 5:** Performance of the classifiers during testing.

| Classifier | Sensitivity | Specificity | Accuracy | AUC-ROC |
| --- | --- | --- | --- | --- |
| Fine Gaussian SVM | 99.6% | 85% | 95.2% | 0.9228 |
| Fine KNN | 73.5% | 98% | 80.9% | 0.8574 |
| Ensemble Bagged Model Trees (EBM) | 87.8% | 97% | 90.6% | 0.9241 |

**Figure 3:**
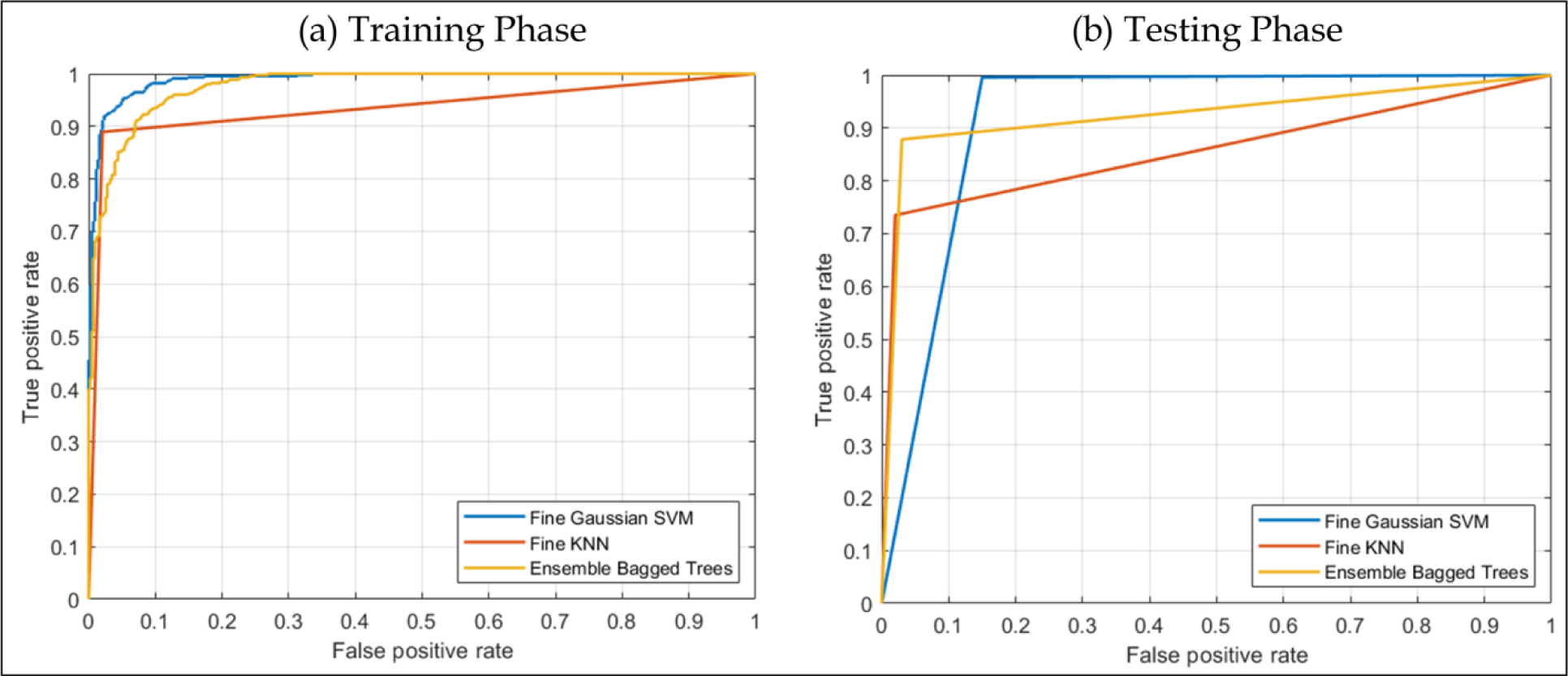
ROC plot of the three best performing classifiers during training (left) and during testing (right).

The ROC curves of all the classifiers during training and test cases are shown in Figure 3.

Table 6 and 7 compare sensitivity and specificity of SVM and EBM classifiers respectively based on the severity. Since the accuracy and AUC-ROC of the fine KNN were less than 90% and 0.9 respectively, it was not considered for further investigation. This is to be noted that for some COVID-19 patients, some of the lung segments were scored as 0 by the radiologists. On the other hand, there were no lung segments with severity score 4 for viral/bacterial pneumonia.

**Table 6:** Sensitivity and specificity based on severity for SVM

|  | Severity | 0 | 1 | 2 | 3 | 4 | Total |
| --- | --- | --- | --- | --- | --- | --- | --- |
| **COVID-19** | Original | 36 | 88 | 71 | 27 | 8 | 230 |
|  | Detected | 35 | 88 | 71 | 27 | 8 | 229 |
|  | Sensitivity | 97.2% | 100.0% | 100.0% | 100.0% | 100.0% | 99.6% |
| **Non-COVID-19** | Original | 50 | 17 | 22 | 11 | 0 | 100 |
|  | Detected | 47 | 11 | 20 | 7 | 0 | 85 |
|  | Specificity | 94.0% | 64.7% | 90.9% | 63.6% | 0.0% | 85.0% |

**Table 7:**
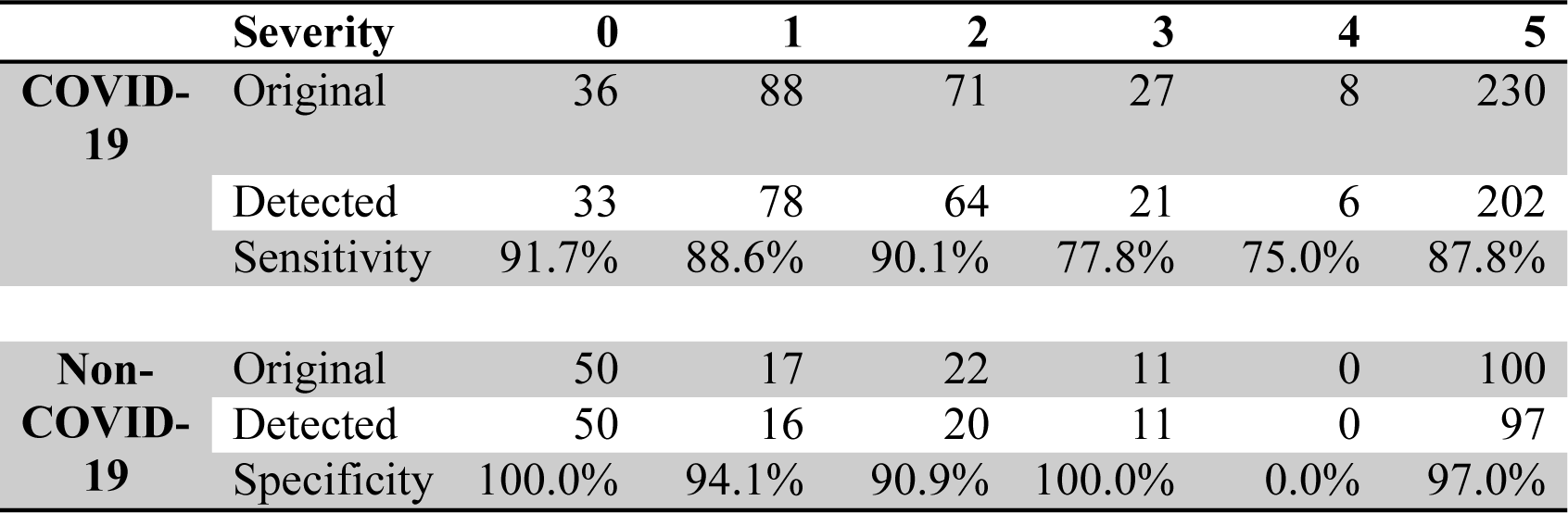
Sensitivity and specificity based on severity for EBM

|  | Severity | 0 | 1 | 2 | 3 | 4 | 5 |
| --- | --- | --- | --- | --- | --- | --- | --- |
| **COVID-19** | Original | 36 | 88 | 71 | 27 | 8 | 230 |
|  | Detected | 33 | 78 | 64 | 21 | 6 | 202 |
|  | Sensitivity | 91.7% | 88.6% | 90.1% | 77.8% | 75.0% | 87.8% |
| **Non-COVID-19** | Original | 50 | 17 | 22 | 11 | 0 | 100 |
|  | Detected | 50 | 16 | 20 | 11 | 0 | 97 |
|  | Specificity | 100.0% | 94.1% | 90.9% | 100.0% | 0.0% | 97.0% |

Sensitivity vs. specificity for both SVM and EBM for the whole test data and at each severity level is plotted in Figure 4.

**Figure 4:**
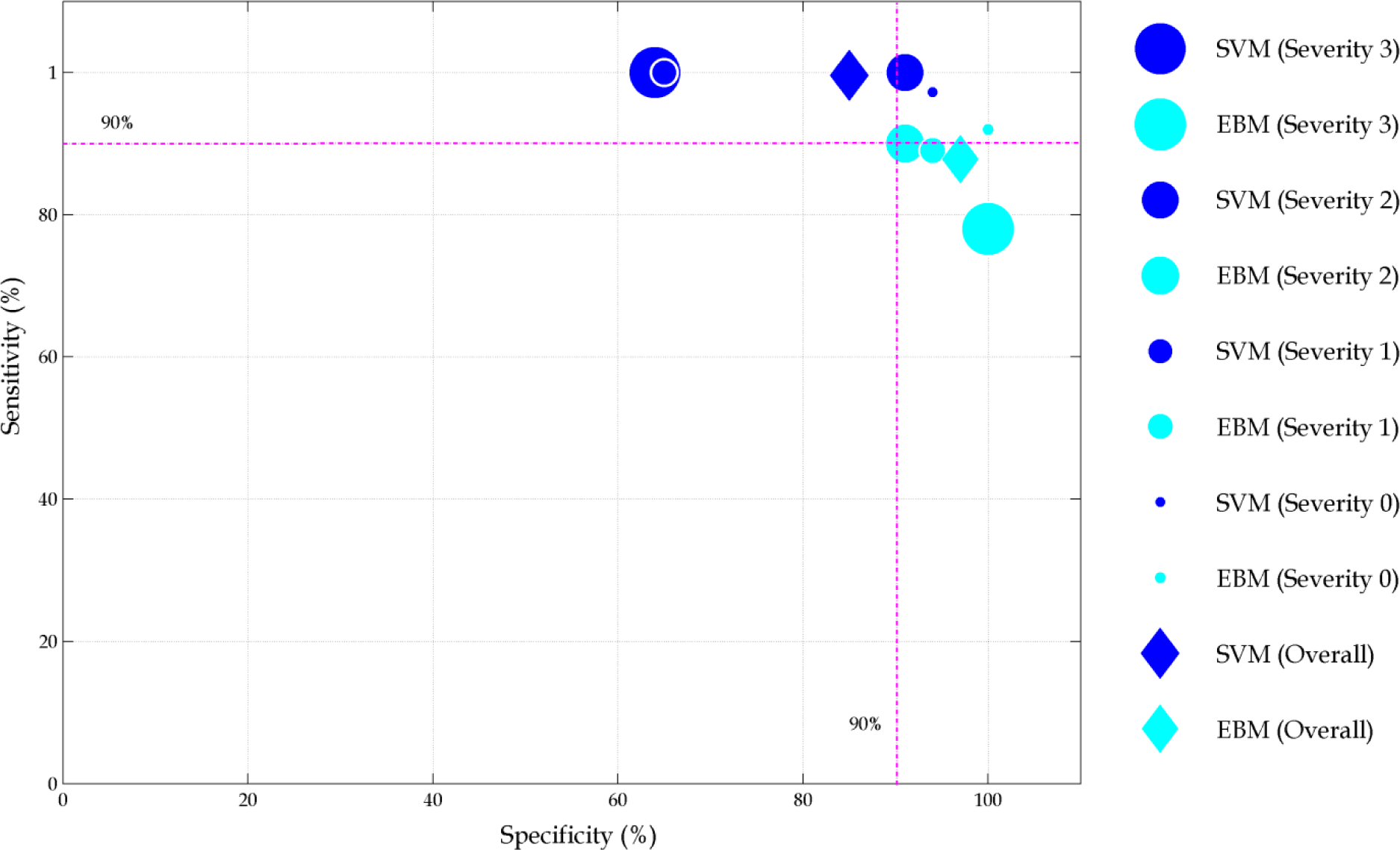
Sensitivity and specificity for whole test data as well as for each severity level. The size of the filled circles represents severity levels from 0 to 4.

Overall, SVM can detect COVID-19 with 99.6% sensitivity and 85% specificity. On the other hand, the performance of EBM is 87.8% and 97% respectively. This implies that there are certain radiomics features that are different for COVID-19 patients. Appropriate selection of those features also can allow to detect COVID-19 from other diseases.

For the whole test data set, the performance of SVM and EBM was comparable. However, KNN performance was much worse compared to the training phase. The reason behind it could be that during training only average sensitivity and specificity of the 10 fold validation were calculated and due to data augmentation via geometric transformation.

The performance between SVM and EBM methods are more distinguishable if severity is taken into consideration. SVM method shows very good sensitivity (97.2 to 100%) but the specificity in terms of severity is not robust with values ranging from 63.6 to 94%. On the other hand, the sensitivity of EBM decreases with the increase of severity. The specificity of the EBM method is within 10% range for all levels of severity and never falls below 90% (range 90.9 to 100%).

## DISCUSSION

It is vital to invent or develop a robust COVID-19 detection tool that can be deployed easily and provide the results within the shortest period time and can be used as a point-of-care screening device. To achieve this goal, a machine learning based approach incorporating radiomics features is proposed in this work that can detect COVID-19 from CXR images. A one-way ANOVA test was used to find the optimal subset of features to train the machine learning classifiers.

Several studies have also been proposed to use different machine learning algorithms to detect COVID-19 from CXR images [10]-[19]-[22]. Most of them have demonstrated good accuracy for disease diagnosis. The performance of those methods vary depending on the definition of the classes used to determine the accuracy. The highest accuracy of 98.08% was achieved by deep learning method [22]. However, it only uses two data classes - COVID-19 and No-Findings. Inclusion of multiple diseases brings the accuracy down to 87.02%. Other studies also reported to achieve similar sensitivity (93 to 96.7%) and specificity (90 to 100%) to classify between COVID-19 and others class, where other class consists of only normal CXR and viral pneumonia. To increase the number of images (sometimes as many as 10 times of the original data), all these algorithm mainly used geometric transformation for data augmentation. Training and test data set were then randomly splited. As a result, it is highly likely that the images of the same patients are present in both the training and test sets resulting in higher accuracies in detection of COVID-19. That is why, it is important to evaluate performance of any machine learning method on a completely independent data representing different lung conditions.

A robust method should be able to detect COVID-19 in presence or absence of other possible lung conditions and the performance should not fluctuate considerably for any other independent data set. In the viewpoint of these criteria, the proposed method is robust as it not only includes normal and viral pneumonia but also other diseases (e.g., bacterial pneumonia, SARS etc.) in the training data as shown in Table 1. The performance is also tested on a completely independent data set. The other uniqueness of this method is that it can provide similar performance irrespective of the severity levels. The authors are not aware of any such studies to investigate image based biomarkers to detect COVID-19 at different severity levels.

Performance evaluation based on severity reveals more insight into the radiomics pattern present in CXR images. Visual assessment of 36 lungs by the radiologists reveals no abnormality and were assigned with the severity score of 0. However, RT-PCR test confirmed COVID-19 positive for these lungs and the proposed method also confirmed COVID-19 positive with a sensitivity of 97.2% and 91.7% by SVM and EBM method respectively.

This work indicates that radiomics features extracted from CXR images can be used as potential image based biomarker to detect and classify COVID-19 in presence of other lung conditions using appropriately selected machine learning algorithm. Considering the performance at different severity levels, EBM method proves to be the most robust method. However, the sensitivity of the EBM method decreases with the increase of severity. One of the reasons could be that with the increase of severity, the patterns that serve as image based biomarkers of COVID-19 vanish and becomes less distinguishable compared to other lung conditions.

## CONCLUSION

The study confirms that CXR images of COVID-19 cases have different patterns than other lung conditions that sometimes are not visible to trained human observer. The study also confirms that with appropriate selection of image based biomarkers (e.g., radiomics features) and machine learning algorithm, it is possible to detect CVOID-19 directly from CXR with a sensitivity and specificity comparable or better than other available techniques, e. g., RT-PCR. The performance of the proposed method with SVM and EBM based machine learning achieved an overall sensitivity of 99.6% and 87.8% and specificity of 85% and 97% respectively. Though the performance are comparable for both the methods, EBM is more robust across severity levels. It will be interesting to investigate the CXR images in cases where RT-PCR results were initially negative and later on was confirmed positive for COVID-19. Since this tool does not require any manual intervention (e.g., sample collection etc.), it can be integrated with any standard X-ray reporting system to be used as an efficient, cost-effective and rapid point-of-care device. It can also be deployed in places where quick results of the COVID-19 test are required, e.g., airport, seaport, hospital, health clinics etc.

## Data Availability

The data related to the findings of this study are available from the authors on reasonable request.

## Code availability

All software code including a single organized script written using MATLAB 2014 will be made available.

## Applicable Funding Source

The authors extend appreciation to the Deputyship for Research & Innovation, Ministry of Saudi Arabia for funding this research work.

