## Supplementary Table 1 for "An Integrated Framework with Machine Learning and Radiomics for Accurate and Rapid Early Diagnosis of COVID-19 from Chest X-ray"

**Supplementary Table 1:** CXR based COVID-19 detection literature

| Name of the paper | Accuracy | sensitivity | specificity | Data Description and Classes | Algorithm Description | Models |
|---|---|---|---|---|---|---|
| **Covallo *et al.*** Texture Analysis in the Evaluation of Covid-19 Pneumonia in Chest X-Ray Images: a Proof of Concept Study | 91.8% | 93% | 90% | positive/negative | After samples separation into training (n = 132) and test (n = 88) sets (60:40 ratio), the training test was used to train 11 classification models: Partial Least Square Discriminant Analysis (PLS-DA), Naïve Bayes (NB), Generalized Linear Model (GLM), Logistic Regression (LR), Fast Large Margin (FML), Deep Learning (DL), Decision Tree (DT), Random Forest (RF), Gradient Boosted Trees (GBT), articial Neural Network (aNN) and Support Vector Machine (SVM) | Ensembl e Machine Learning (cut-off 132.57) |
| **Asif *et al.*** Classification of COVID-19 from Chest X-ray images using Deep Convolutional Neural Networks Sohaib Asif, Yi Wenhui*, Hou Jin, Yi Tao, | 98% (training accuracy of 97% and validation accuracy of 93%). | NA | NA | Normal 1341, Viral Pneumonia 1345, COVID-19 864 (data augmentation and splitting for training, validation and test) | 88% for training, 7% for validation and 5% for testing | based model Inception V3 with transfer learning have been CNN |
| **Basu *et al.*** Deep Learning for Screening COVID-19 using Chest X-Ray Images | 95.3% ± 0.02 overall,100% of the covid and normal cases being correctly classified in each validation fold | -NA | NA | Normal, Pneumonia, COVID-1, other disease (atelectasis, cardiomegaly, infiltration, effusion, nodule, mass)<br><br>225 Covid-19 108,948 frontal chest x-ray<br><br>5-fold cross validation | The adopted procedure is outlined below.<br>• Instantiate the convolutional base of the model trained on Data-A and load its pre-trained weights. • Replace the last fully connected layer of the pre-trained CNN with a new fully connected layer.<br>• Freeze the layers of the model up to the last convolutional block. • Finally retrain the last convolution block and the fully connected layers using Stochastic Gradient Descent (SGD) optimization algorithm with a very slow learning rate. | CNN, Connected with Gradient Class Activation Map (Grad-CAM) for detecting the regions<br><br>Domain extension transfer learning (DETL) |
| **Ahmed 2020 *et al*** ReCoNet - Multi-level Preprocessing of Chest X-rays for COVID-19 Detection Using Convolutional Neural Networks | 97.48% | 96.39% | 97.53% | **Training:**<br>COVID-19 207, Normal 7966, Pneumonia 5451<br>**Test:**<br>COVID-19 31, Normal 885, Pneumonia 594 | Used 90% of the data for training and 10% for validation. | ReCoNet – preprocessing before feeding to deep net in the same framework |
| **Chowdhury 2020** Can AI help in screening Viral and COVID-19 pneumonia | Normal vs Covid-19 98.3%<br><br>Normal vs. viral vs. Covid-19 98.3% | Normal vs Covid-19 96.7%<br><br>Normal vs viral vs Covid-19 96.7% | Normal vs Covid-19 100%<br><br>Normal vs viral vs Covid-19 99% | **Training:**<br>COVID-19 130, Normal 190, Viral Pneumonia 190<br>**Test:**<br>COVID-19 60, Normal 1151, Viral Pneumonia | | Four different network: 1) ALexNet, 2) ResNet28, 3) DenseNet201 and 4) SqueezeNet |

| | | | | | | |
|---|---|---|---|---|---|---|
| | | | | 594 | | |
| **Ozturk *et al.*** Automated detection of COVID-19 cases using deep neural networks with X-ray images | 98.08% | 95.3 | 95.3 | binaryclass classification (COVID vs. No-Findings | 80% of X-ray images are used for training and 20% for validation. | Dark CovidNet architecture |
| | 87.02% | 85.35 | 92.18 | Multiclass (COVID vs. No-Findings vs Pneumomia) | | |
