## Supplementary Table 2 for "An Integrated Framework with Machine Learning and Radiomics for Accurate and Rapid Early Diagnosis of COVID-19 from Chest X-ray"

**Supplementary Table 2:** p-value of the significant features extracted using ANOVA

| Feature No. | Feature Name | p |
|---|---|---|
| 1 | **Firstorder 10 Percentile** | 0.045053 |
| 2 | **Firstorder 90 Percentile** | 1.73E-05 |
| 3 | **Firstorder Entropy** | 0.00967319 |
| 4 | **Firstorder Interquartile Range** | 0.00146048 |
| 5 | **Firstorder Maximum** | 1.06E-10 |
| 6 | **Firstorder Mean** | 0.01466991 |
| 7 | **Firstorder Mean Absolute Deviation** | 0.00119999 |
| 8 | **Firstorder Minimum** | 0.02584477 |
| 9 | **Firstorder Range** | 5.79E-08 |
| 10 | **Firstorder Robust Mean Absolute Deviation** | 0.00123508 |
| 11 | **Firstorder Root Mean Squared** | 0.00452603 |
| 12 | **Firstorder Skewness** | 1.06E-10 |
| 13 | **Firstorder Variance** | 0.00027519 |
| 14 | **GLCM Cluster Prominence** | 0.00019394 |
| 15 | **GLCM Cluster Shade** | 1.23E-09 |
| 16 | **GLCM Cluster Tendency** | 0.00029227 |
| 17 | **GLCM Contrast** | 5.41E-10 |
| 18 | **GLCM Correlation** | 1.24E-05 |
| 19 | **GLCM Difference Average** | 1.55E-10 |
| 20 | **GLCM Difference Entropy** | 1.07E-10 |
| 21 | **GLCM Difference Variance** | 1.11E-10 |
| 22 | **GLCM Id** | 1.36E-10 |
| 23 | **GLCM Idm** | 1.43E-10 |
| 24 | **GLCM Idmn** | 5.58E-07 |
| 25 | **GLCM Idn** | 4.70E-10 |
| 26 | **GLCM Imc1** | 1.73E-10 |
| 27 | **GLCM Imc2** | 0.00092835 |
| 28 | **GLCM Inverse Variance** | 1.24E-10 |
| 29 | **GLCM Joint Energy** | 0.00063385 |
| 30 | **GLCM Joint Entropy** | 3.64E-07 |
| 31 | **GLCM Maximum Probability** | 0.00496733 |
| 32 | **GLCM Sum Entropy** | 8.69E-06 |
| 33 | **GLCM Sum Squares** | 0.00018272 |
| 34 | **GLDM Dependence Entropy** | 2.14E-08 |
| 35 | **GLDM Dependence Non Uniformity Normalized** | 2.95E-10 |
| 36 | **GLDM Dependence Variance** | 0.00193179 |
| 37 | **GLDM Gray Level Variance** | 0.0002227 |
| 38 | **GLDM Large Dependence Emphasis** | 1.16E-09 |
| 39 | **GLDM Low Gray Level Emphasis** | 0.02110404 |
| 40 | **GLDM Small Dependence Emphasis** | 0.00349533 |
| 41 | **GLDM Small Dependence High Gray Level Emphasis** | 0.0017444 |
| 42 | **GLRLM Gray Level Variance** | 0.00084841 |
| 43 | **GLRLM High Gray Level Run Emphasis** | 0.00143554 |
| 44 | **GLRLM Long Run Emphasis** | 3.97E-06 |

| | | |
|---|---|---|
| 45 | **GLRLM Long Run High Gray Level Emphasis** | 8.93E-06 |
| 46 | **GLRLM Low Gray Level Run Emphasis** | 0.00189904 |
| 47 | **GLRLM Run Percentage** | 6.57E-09 |
| 48 | **GLRLM Run Variance** | 1.84E-06 |
| 49 | **GLRLM Short Run Emphasis** | 5.90E-06 |
| 50 | **GLRLM Short Run High Gray Level Emphasis** | 0.00071359 |
| 51 | **GLRLM Short Run Low Gray Level Emphasis** | 1.06E-10 |
| 52 | **GLSZM Gray Level Non Uniformity Normalized** | 1.25E-10 |
| 53 | **GLSZM Gray Level Variance** | 6.25E-09 |
| 54 | **GLSZM High Gray Level Zone Emphasis** | 0.00035915 |
| 55 | **GLSZM Large Area Emphasis** | 0.00239442 |
| 56 | **GLSZM Large Area High Gray Level Emphasis** | 0.01690471 |
| 57 | **GLSZM Large Area Low Gray Level Emphasis** | 0.00048366 |
| 58 | **GLSZM Low Gray Level Zone Emphasis** | 1.23E-05 |
| 59 | **GLSZM Size Zone Non Uniformity Normalized** | 1.06E-10 |
| 60 | **GLSZM Small Area Emphasis** | 1.06E-10 |
| 61 | **GLSZM SmallAreaLowGrayLevelEmphasis** | 1.06E-10 |
| 62 | **GLSZM Zone Entropy** | 1.06E-10 |
| 63 | **GLSZM Zone Percentage** | 0.02236129 |
| 64 | **GLSZM Zone Variance** | 0.00243722 |
| 65 | **NGTDM Coarseness** | 4.01E-06 |
| 66 | **NGTDM Complexity** | 3.41E-08 |
| 67 | **NGTDM Contrast** | 9.77E-09 |
| 68 | **NGTDM Strength** | 1.45E-10 |
| 69 | **Shape2D Elongation** | 1.23E-07 |
| 70 | **Shape2D Perimeter** | 1.30E-10 |
| 71 | **Shape2D Sphericity** | 1.06E-10 |
